# What does it mean to use the mean? The impact of different data handling strategies on the proportion of children classified as meeting 24-hr movement guidelines and associations with overweight and obesity

**DOI:** 10.1101/2023.09.22.23295801

**Authors:** Christopher D. Pfledderer, Sarah Burkart, Roddrick Dugger, Hannah Parker, Lauren von Klinggraeff, Anthony D. Okely, R. Glenn Weaver, Michael W. Beets

## Abstract

**Background:** Despite the widespread endorsement of 24-hour movement guidelines (physical activity, sleep, screentime) for youth, no standardized processes for categorizing guideline achievement exists. The purpose of this study was to illustrate the impact of different data handling strategies on the proportion of children meeting 24-hour movement guidelines (24hrG) and associations with overweight and obesity.

*Methods:* A subset of 524 children (ages 5-12yrs) with complete 24-hour behavior measures on at least 10 days was used to compare the impact of data handling strategies on estimates of meeting 24hrG. Physical activity and sleep were measured via accelerometry. Screentime was measured via parent self-report. Comparison of meeting 24hrG were made using 1) average of behaviors across all days (AVG-24hr), 2) classifying each day and evaluating the percentage meeting 24hrG from 10-100% of their measured days (DAYS-24hr), and 3) the average of a random sample of 4 days across 10 iterations (RAND-24hr). A second subset of children (N=475) with height and weight data was used to explore the influence of each data handling strategy on children meeting guidelines and the odds of overweight/obesity via logistic regression.

*Results:* Classification for AVG-24hr resulted in 14.7% of participants meeting 24hrG. Classification for DAYS-24hr resulted in 63.5% meeting 24hrG on 10% of measured days with <1% meeting 24hrG on 100% of days. Classification for RAND-24hr resulted in 15.9% of participants meeting 24hrG. Across 10 iterations, 63.6% of participants never met 24hrG regardless of the days sampled, 3.4% always met 24hrG, with the remaining 33.0% classified as meeting 24hrG for at least one of the 10 random iterations of days. Using AVG-24hr as a strategy, meeting all three guidelines associated with lower odds of having overweight obesity (OR=0.38, p<0.05). The RAND-24hr strategy produced a range of odds from 0.27 to 0.56. Using the criteria of needing to meet 24hrG on 100% of days, meeting all three guidelines associated with the lowest odds of having overweight and obesity as well (OR=0.04, p<0.05).

*Conclusions:* Varying estimates of meeting the 24hrG and the odds of overweight and obesity results from different data handling strategies and days sampled.

## BACKGROUND

The 24-hour movement guidelines (24hrG) for children outline an optimal composition of movement behaviors for the 24-hour day. The guidelines recommend 60 minutes per day of moderate to vigorous physical activity (MVPA), 9-11 hours of sleep per night for children aged 5-13 years, and no more than 2 hours per day of recreational screentime.^1^ The guidelines have been adopted by multiple countries and agencies over the past decade,^2–5^ and researchers have followed suit by integrating the movement guidelines in analyses for observational and intervention research.^6^ Achieving the 24hrG is linked with multiple health-related outcomes among youth, including adiposity,^7^ fitness,^8^ health-related quality of life,^9,10^ mental, emotional, and social health,^11,12^ dietary patterns,^13^ social-cognitive development,^14,15^ and bone and skeletal health.^16^ Despite the widespread endorsement of the 24hrG, there are no standardized ways to process movement data prior to classifying participants as meeting or not meeting guidelines or associating guideline adherence with health outcomes. A variety of methods are used to do so, including averaging across the total measurement period and using daily estimates of movement behaviors to quantity adherence to the 24hrG.^17^

Common practice in handling movement behavior data is to average behaviors across multiple days (typically a minimum of three weekdays and one weekend day).^17^ The 24hrG, however, are framed as meeting recommended amounts of all three behaviors “each day”, which implies the 24hrG should be met on each individual day. Using estimates averaged across days allows for children to have some “days off” where they do not meet a 24hrG. However, studies have demonstrated natural day-to-day variability in youth movement behavior patterns,^18–24^ with some days, like school or weekends, associated with higher or lower levels of a movement behavior. For example, in a four-day window comprised of two weekdays and two weekend days, a child may be relatively inactive and not meet the physical activity guidelines during the weekend where they accumulate 30min/day MVPA but participate in sports during the week where they engage in 90min/day MVPA. Taking each day separately, this would result in meeting the MVPA guideline on two of the four days. Conversely, a second child may accumulate 90 minutes of MVPA on one day and 40 minutes of MVPA on the other three days. Using the average across all days, the first child would be classified as meeting the physical activity guideline, but only met the guideline on half of the days, whereas the second child would be classified as not meeting the guideline yet would have eclipsed the guideline on one of four days.

While the impact of day-to-day variability on estimates of meeting 24hrG is unknown, it raises important questions as to how best to process multi-day movement behavior data to estimate the prevalence of children meeting the guidelines and subsequent associations with overweight and obesity. Understanding the impact of different data handling procedures of multi-day movement behaviors in the context of evaluating 24hrG adherence, health outcomes, and the natural day-to-day variability in youth movement behaviors is important because each method could lead to an under/over-estimation of meeting the guidelines, which may influence the relationship between meeting guidelines and health outcomes and obscure the relationship between predictors of meeting guidelines.

The purpose of this study was to illustrate the day-to-day variability of movement behaviors in a large cohort of children and show the differences among three data handling strategies on the estimated proportion of children meeting the 24hrG and associations with overweight and obesity: 1) using averages derived from total observed days, 2) evaluating the percentage of children meeting the guidelines from 10-100% of days measured, and 3) using averages derived from a random sample of four days (three weekdays and one weekend day) across 10 iterations.

## METHODS

### Data Collection

Data came from an ongoing longitudinal cohort study which measured children’s movement behaviors (i.e., physical activity, sedentary time, sleep, screentime during Fall 2020 during school (Oct/Nov), Spring 2021 during school (April/May), and Summer 2021 when school was out (July). It is worth noting, during Fall 2020, participants experienced a hybrid learning situation in which school was delivered virtually three days/week and was attended in-person two days/week. In-person school resumed fulltime in the Spring of 2021. Due to the COVID-19 pandemic, limited height and weight data (used to quantify the prevalence of overweight and obesity) was collected. For the purpose of understanding how different data handling strategies would influence associations with overweight and obesity, we utilized data collected in Spring 2022 during school (April/May), which had complete height and weight data for children in the study. Participants were recruited via two neighboring school districts in the southeastern United States which served K-6^th^ grades between January-April 2021 and during January-April 2022. No exclusion criteria were used prior to recruitment. All procedures were approved by the university’s Institutional Review Board prior to the start of the study (IRB#Pro00080382) and participant consent was obtained prior to being enrolled in the study. Authors had access to identifiable participant information at the beginning of data collection to distribute accelerometers via mail to participants and after data collection to ensure appropriate linkages of data across sources (accelerometry, surveys, and height/weight measurements.

Device-measured MVPA, device-measured measured sleep, and daily collection of parent-reported children’s screentime were utilized for this study. Within each data collection period, children were asked to wear an Actigraph GT9X accelerometer on their non-dominant wrist 24 hours per day for 14 days. Additionally, parents received a brief online survey (daily diary) each night of the 14-day wear period and were asked to provide information about their child’s day, including screentime and bed and wake time. After the 14-day wear period, accelerometers were downloaded, and data were prepared for processing. Screentime was assessed with two items on the parent-completed daily diary. First, parents were asked if their child watched a screen at home today (e.g., watch TV, play video games, used a smartphone, used a tablet, laptop, or desktop computer). If parents responded, “Yes,” then they were prompted to estimate the total time spent watching a screen from a dropdown list in 30-minute increments up to 10 hours. The daily screen time estimate was calculated by adding reported time spent across all devices. The decision to assess screen time in increments of 30 minutes was two-fold. First, because the survey question for screen time was provided in a drop-down format in Qualtrics, this option only allowed for a restricted number of response options available to the end-user (participants). Second, other large-scale observational studies, such as NHANES, “bin” response options into larger time units on the backend when analyzing outcomes and presenting results. Further, for studies that use instruments (IPAQ, NHANES) that ask for the number of minutes (in one-minute increments) the vast majority of responses by adults are in 30-minute or longer increments. Thus, while we recognize the limitations of collecting our screen time data in 30-minute increments, the evidence suggests even if participants are provided with an option to respond to the nearest minute, they typically “self bin” their responses into larger time increments, such as 30 minutes.

Height (cm) and weight (kg) were measured using standard procedures (digital scale to the nearest 0.01kg [Healthometer model 500KL, Health o meter, McCook, IL], stadiometer to nearest 0.1cm [Model S100; Aytron Corp., Prior Lake, MN], without shoes, wearing light clothing) by trained male and female research assistants using discrete procedures (e.g., females measure females, behind changing screen). During school, height and weight were measured at the beginning of the day using the Physical Education classroom (e.g., gymnasium). Body Mass Index (BMI) was calculated (BMI = kg/m^2^) and transformed into age- and sex-specific BMI z-scores. Overweight and obesity was defined as at or above the 85^th^ percentile.

### Data Processing

#### Accelerometry

Actigraph GT9X accelerometers were initialized and downloaded using Actilife software (version 6.13.4, Pensacola, FL). Accelerometers were initialized to record data at a frequency of 30 Hz and began data collection at 7:00 AM on the day preceding earliest device delivery. Stop time was not used. Idle sleep mode was enabled to preserve battery life and the display was turned off to limit distractions for children while attending school. Data were downloaded and saved in raw format as GT3X files and converted to .csv files for processing. Raw .csv files were processed using the GGIR package (version 2.6-0)^25^ in R (Version 4.1.2; R Foundation for Statistical Computing; Vienna, Austria). Time spent in physical activity intensity categories was determined using intensity thresholds described by Hildebrand et al.^26^ Sleep estimates were guided by use of parent-reported bed and wake times. On nights when parents provided bed and wake times, the advanced sleep log option in GGIR was used to guide detection of the sleep period and when bed and wake times were not provided, GGIR’s HDCZA algorithm was used to detect the sleep period.^27^

### Inclusion and Exclusion Criteria for Valid Data

Because there are 24hrG written specifically for children aged 5-12 years, participants were excluded if they were ≤ 4 years old. Participants must have provided valid accelerometry data (at least 16 hours of wear time per day) for both physical activity and sleep, and parent reports of screentime, on at least 10 days for at least one timepoint (Fall 2020, Spring 2021, or Summer 2021). For the second part of the study, where associations with overweight and obesity were explored, inclusion criteria for movement behaviors was the same, and in addition, participants must have complete height and weight data used to calculate BMI and BMI z-scores.

### 24-Hour Movement Behavior Criteria

For all analyses, participants were considered to have met guidelines based on the following criteria:

- Physical activity: An accumulation of ≥ 60 minutes per day of MVPA
- Sleep: 9-11 hours of sleep per night
- Screentime: ≤ 2 hours per day of recreational screentime

### Analysis of 24hr Movement Behaviors and Variability

Physical activity, sleep, and screentime were summarized descriptively for the total sample. In addition to descriptive summaries, day-to-day variability of movement behaviors was illustrated in a random sample of four participants who all had valid data and who were measured on the exact same calendar dates across 10 days. Daily values of each movement behavior for each of these participants are presented as well as their average values across the 10-day window.

### Data Handling Strategies

Table 1 summarizes each data handling strategy, including a general description of the data structure and how the results are presented.

**Table 1.** Summary of data handling strategies.

| Data Handling Strategy | Data Description | Description of Strategy | Data Presentation |
| --- | --- | --- | --- |
| AVG-24hr | Data from participants who had at least 10 valid days of physical activity, sleep, and screentime data | Daily estimates were averaged.<br>Averages were dichotomized into “met guidelines” and “did not meet guidelines” for the total sample | The percentage of participants meeting the guidelines for each behavior, separately, and for all three behaviors for the total sample |
| DAYS-24hr | Data from participants who had at least 10 valid days of physical activity, sleep, and screentime data | Each individual day was dichotomized as “met” or “did not meet” a guideline | The percentage of participants meeting the guidelines at each criterion, starting at 10% (e.g., 1 of 10 days) of days and going up to 100% (e.g., 10 of 10 days) of their days |
| RAND-24hr | Four days (three weekdays, one weekend day) of data randomly selected from the total days of valid data from each participant | Random sampling of four days was repeated over 10 rounds and the total number of times participants met guidelines using those four days of data was calculated for each round of randomization | 1) The percentage of participants meeting the guidelines for each behavior for each random sample<br>2) The total number of times participants met each guideline across the 10 rounds of randomization, ranging from 0 to 10 |

#### Data Handling Strategy #1 – Averaging Total Days of Data (AVG-24hr)

Strategy #1 utilized all days of valid data from included participants. Consistent with current practices, a single average was calculated for each movement behavior across all available days. Children were classified as meeting or not meeting each of the three guidelines based on the average. Results are presented as the percent of participants meeting each movement behavior guideline and the percentage meeting all three guidelines.

#### Data Handling Strategy #2 – “Percent Days” Criteria (DAYS-24hr)

Strategy #2 also utilized all days of valid data from included participants. Participants were classified as meeting or not meeting the guidelines based on the percent of individual days on which they met the guideline. The behaviors on each day were classified as meeting or not meeting the guideline. The total number of days meeting each individual guideline and all three guidelines was calculated and divided by the total number of days of valid data (i.e., minimum of 10 days). The percentage of days meeting the guidelines was calculated. Results for this strategy are presented as the percentage of participants meeting guidelines for each of the percent days necessary to meet the guidelines.

#### Data Handling Strategy #3 – Random Sampling of Four Days (RAND-24hr)

Strategy #3 created average values from random samples of children’s data using four valid days (3 weekdays and 1 weekend day). Averaging movement behaviors across four days is consistent with current practice in the literature.^17^ To do this, four days (three weekdays and one weekend day) of data were randomly selected from the total valid days for each participant. This randomization process was repeated over 10 iterations using STATA’s ‘rannum’ command and the ‘set seed’ feature. Averages were calculated for each movement behavior for each participant for each of the 10 random samples of four days. Results are presented as the percent of participants meeting each movement behavior guideline consistent to Strategy #1-AVG-24hr. A second outcome from Strategy #3 was also calculated that represented the total number of times participants met each guideline across the 10 iterations of randomization, ranging from 0 times (participants never met the guideline regardless of days sampled across all 10 iterations of randomization) to 10 times (participants always met the guidelines, regardless of the days sampled).

### Associations with Overweight and Obesity

The odds of overweight and obesity across different data handling strategies were derived from logistic regression models, with overweight and obesity used as the binary outcome variable (0= below 85^th^ percentile, 1 = above 85^th^ percentile), guideline adherence as either the binary predictor variable (0 = did not meet guideline, 1 = met guideline) or continuous based on the percentage of days a guideline was met, and covariates including sex, race/ethnicity, age, and household income. Logistic regression models were constructed for each of the data handling strategies and for each of the movement behaviors, including meeting all three guidelines. Marginal predicted probabilities of overweight and obesity were also calculated based on logistic regression models.

## RESULTS

### Data and Participant Characteristics

Table 2 shows the descriptive data and participant characteristics for each analytical sample. For guideline adherence, a total of 524 participants (K-6^th^ grade [mean age = 8.6 ± 1.7 years], 49% female, 66% White) were included in the analyses. Participants (N=524) averaged 68.9 ± 28.1 minutes/day of MVPA, 9.1 ± 0.7 hours/day of sleep, and 182.9 ± 116.1 minutes/day of screentime. For guideline adherence and subsequent associations with overweight and obesity, a total of 475 participants (K-6^th^ grade [mean age = 9.5 ± 1.8 years], 49% female, 55% White) were included in the analyses. Participants (N=475) averaged 67.1 ± 28.1 minutes/day of MVPA, 8.9 ± 0.8 hours/day of sleep, and 128.5 ± 97.0 minutes/day of screentime.

**Table 2.**
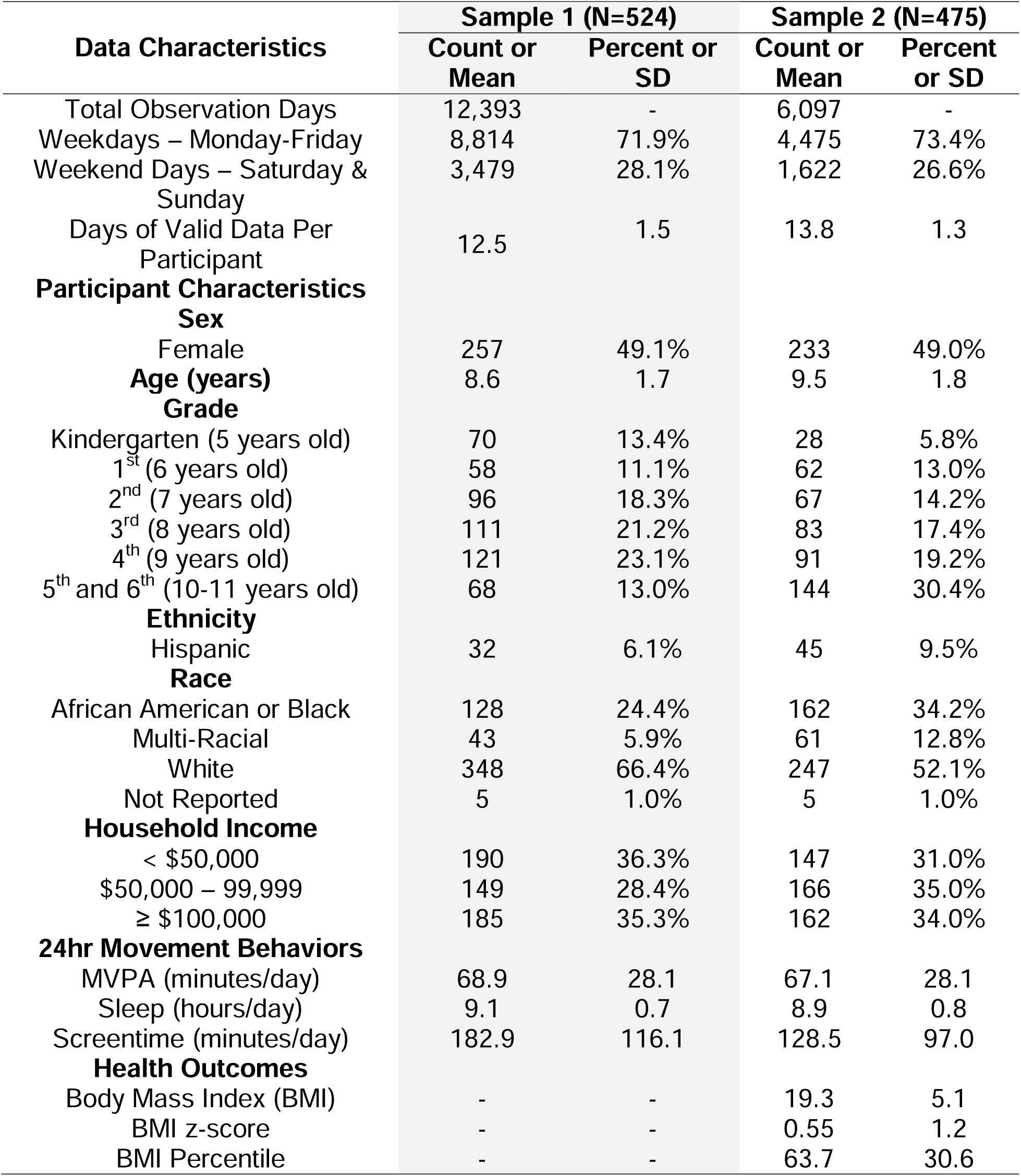
Data and participant characteristics.

### Summary of 24hr Movement Behavior Variability

Figures 1 and 2 illustrate the day-to-day variability in the sample for each of the individual movement behaviors. Figure 1 shows box plots of each movement behavior for each individual in the sample. Individual box plots are color coded to communicate participants who meet (orange) and did not meet (blue) each guideline based on their total average across all measurement days (the most common way of handing multi-day data) and are sorted (left to right) based on the average. The boxplots visually summarize the average across all measured days, with error bars indicating standard deviation from the mean. These plots illustrate that all participants, whether they meet or do not meet the guidelines on average, meet or exceed the behavioral guideline threshold (indicated as a solid black line) on some of the days monitored and not on others. Figure 2 highlights the day-to-day variability of these movement behaviors for a random sample of four participants, all who had valid data and were assessed on the exact same dates for 10 days. Using averages, one participant (Participant #2) met MVPA guidelines, and two participants (Participant #2 and Participant #3) met sleep and screentime guidelines. On the day-level, there was considerable variability in movement behaviors across the 10-day period.

**Figure 1.**
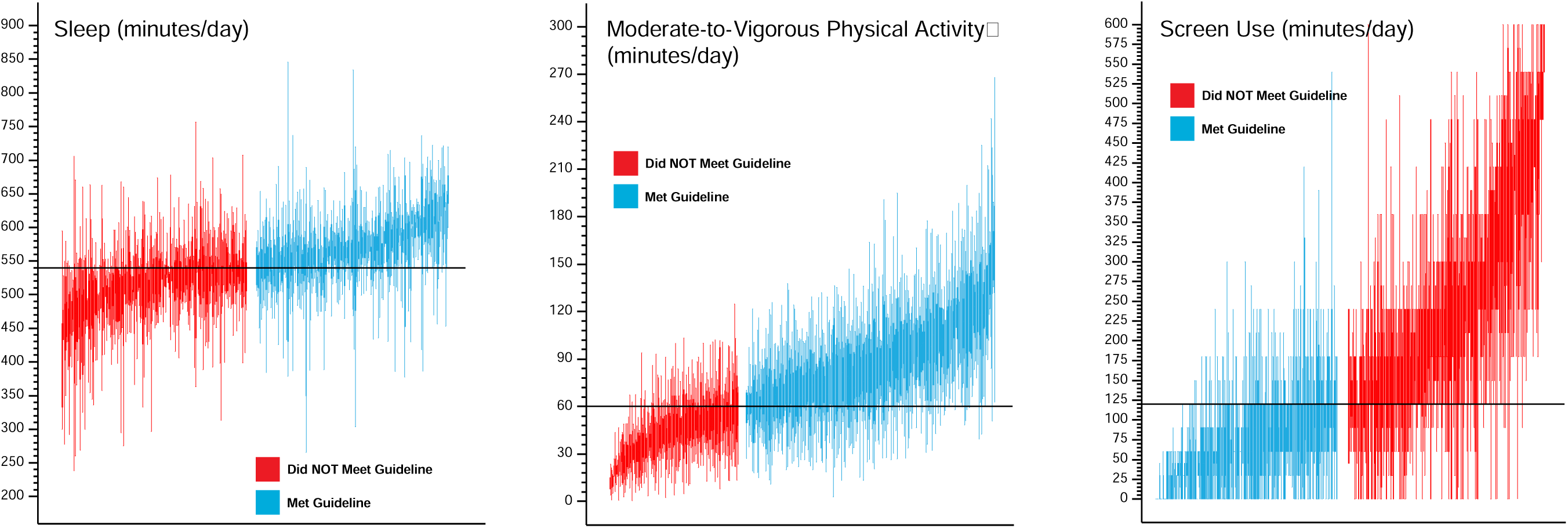
Boxplots of each movement behavior for each individual in the total sample (N=524).

**Figure 2.**
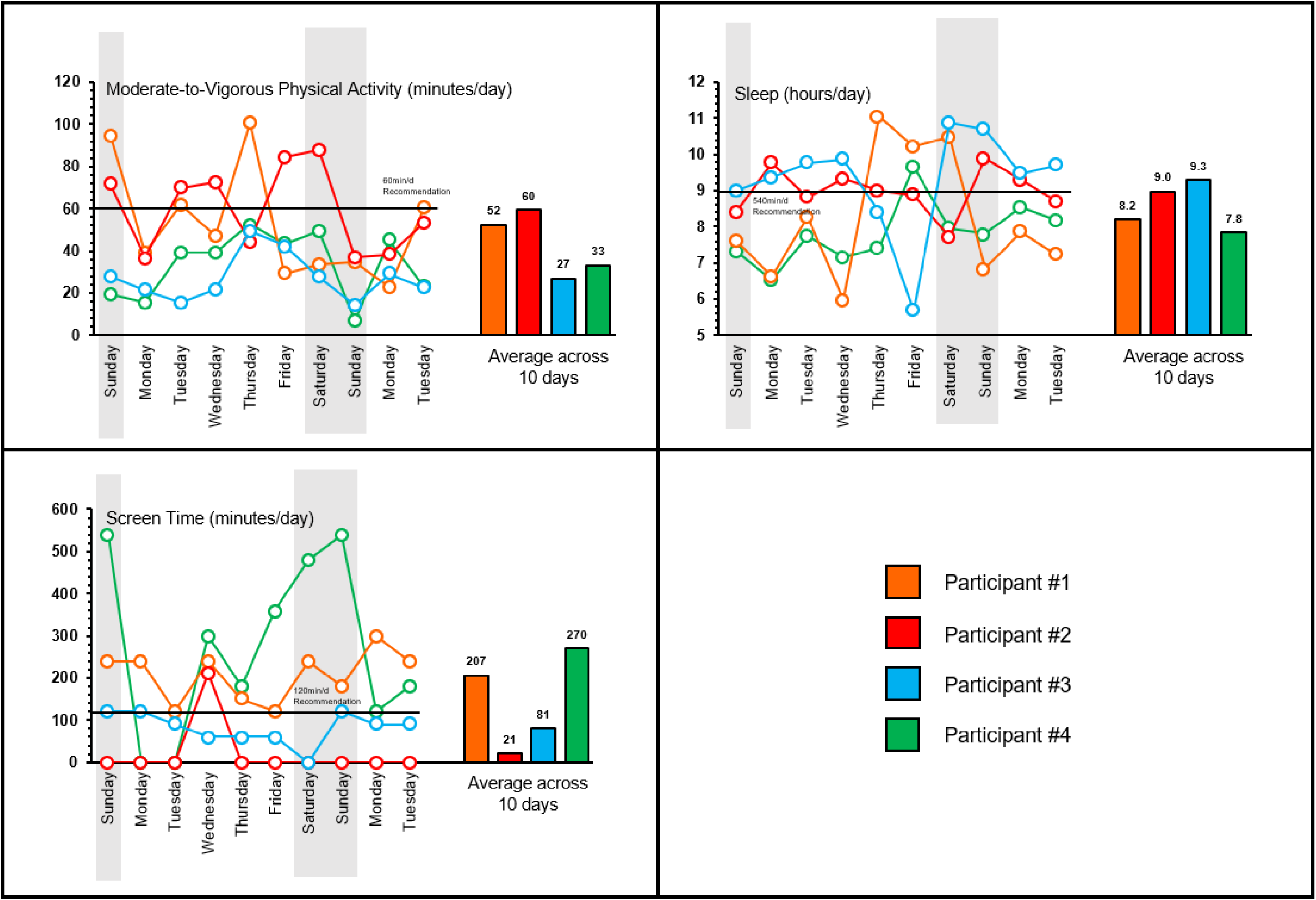
Variability of movement behaviors across 10 days from a randomly selected sample of four participants.

### Data Handling Strategy #1 – Averaging Total Days of Data (AVG-24hr)

Figure 3 displays the percentage of participants meeting each movement guideline based on the average of their total observed days (dotted horizontal lines). MVPA guidelines were met by 59.4% of participants, sleep guidelines were met by 54.9% of participants, screentime guidelines were met by 33.2% of participants, and 14.7% of participants met all three guidelines. Figure 4 displays the odds of overweight and obesity for participants meeting guidelines using the average of their total observed days (red circle). Participants meeting MVPA (OR=0.33, p<0.05), Sleep (OR=0.56, p<0.05), Screentime (OR=0.83, p<0.05), and all three 24hrG (OR=0.38, p<0.05) had lower odds of overweight/obesity using theAVG-24hr data handling strategy.

**Figure 3.**
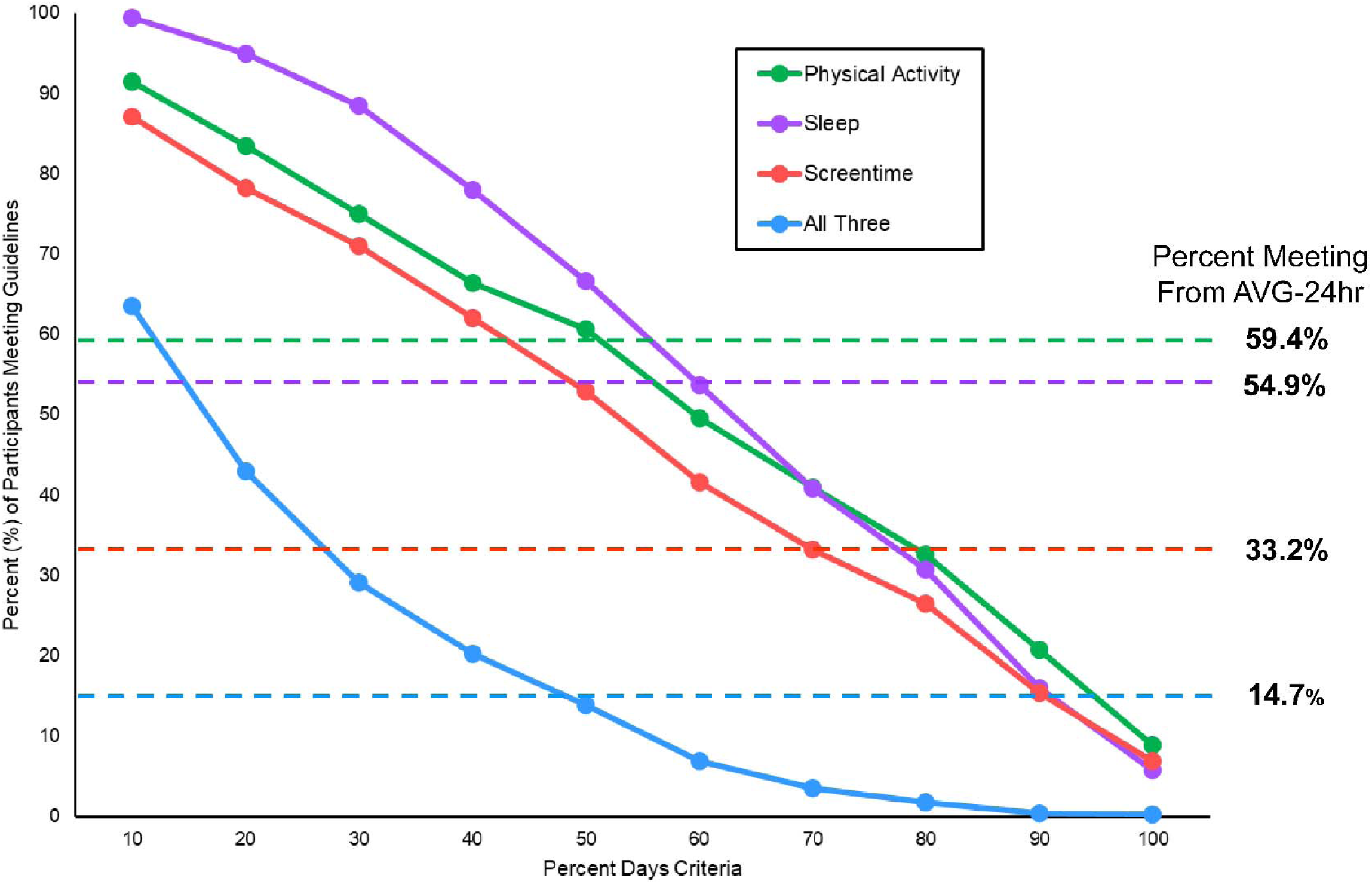
Prevalence of meeting movement guidelines based on “percent days” criteria (solid lines) compared to the percent of participants meeting guidelines based on averaging total days of data (dotted horizontal lines).

**Figure 4.**
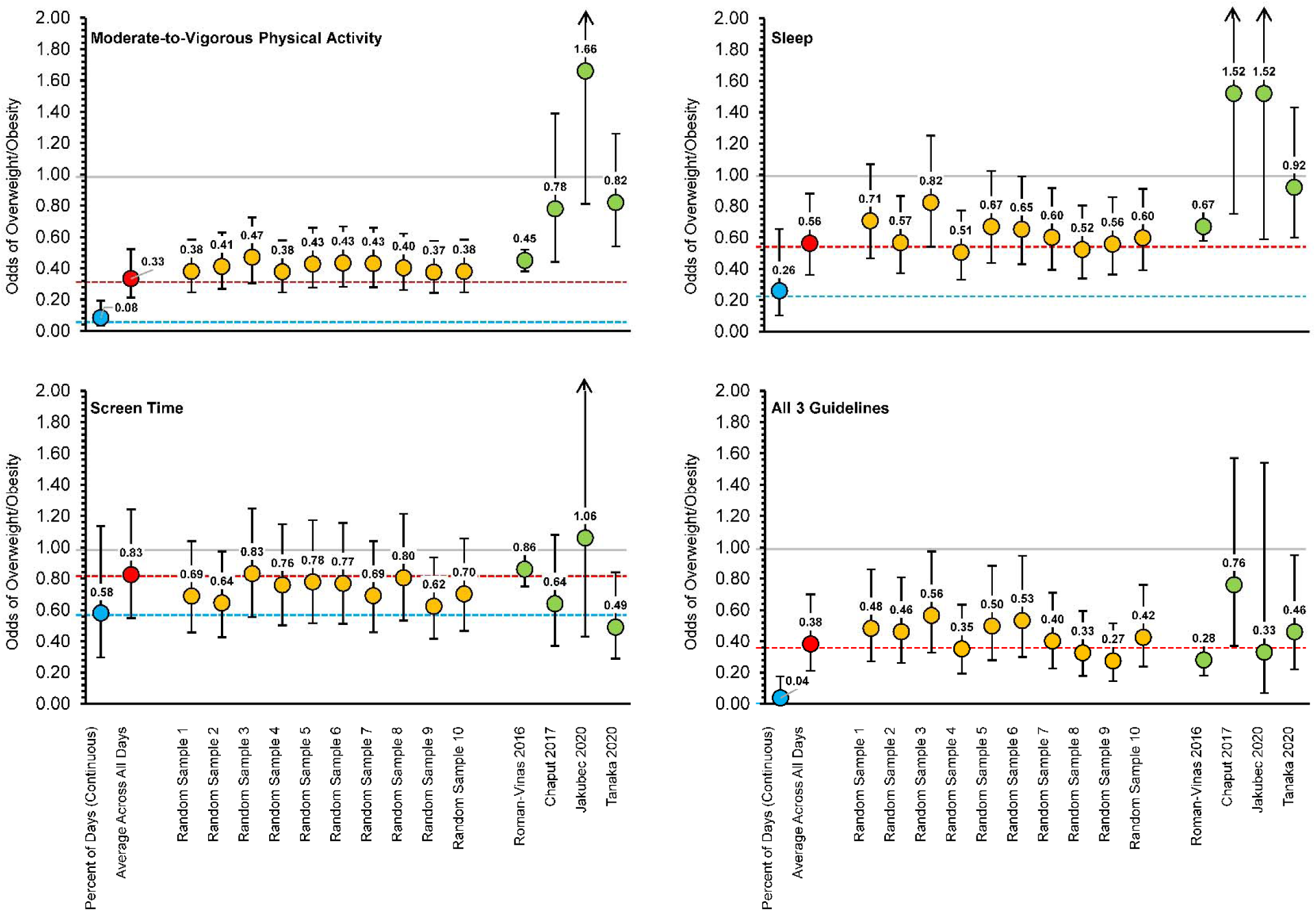
Odds of overweight and obesity across different data handling strategies and compared to previously published findings

### Data Handling Strategy #2 – “Percent Days” Criteria (DAYS-24hr)

The prevalence of meeting movement guidelines based on DAYS-24hr is presented in Table 3 and visually illustrated in Figure 3 (solid lines). When using the criteria of only needing to meet guidelines on 10% of observed days (e.g., 1 out of 10 days), 91.5% of participants met the physical activity guidelines, 99.4% met sleep guidelines, 87.1% met screentime guidelines, and 63.5% met all three guidelines. When using the criteria of needing to meet guidelines on at least 50% of observed days (e.g., 5 out of 10 days), 60.6% of participants met MVPA guidelines, 66.6% met sleep guidelines, 53.0% met screentime guidelines, and 13.9% met all three guidelines. When using the criteria of needing to meet guidelines on 100% of observed days (i.e., meet guidelines every day), 8.9% of participants met MVPA guidelines, 5.7% met sleep guidelines, 6.9% met screentime guidelines, and 0.3% met all three guidelines. The predicted probabilities of overweight and obesity for each participant across the percentage of days a guideline was met is displayed in Figure 5. There was a clear downward trend in the predicted probability of overweight and obesity as the percentage of days participants met guidelines increased from 10% of days to 100% of measured days.

**Figure 5.**
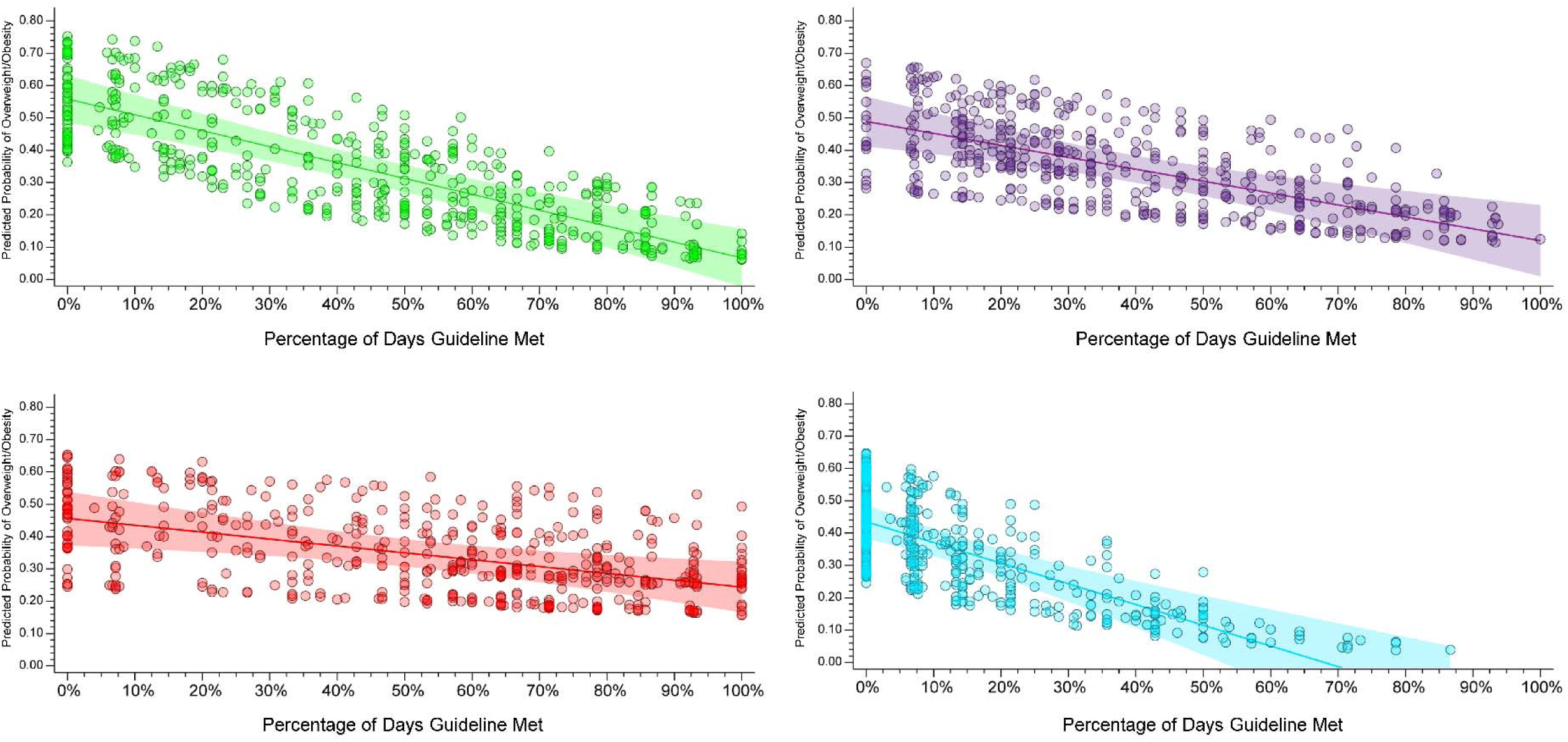
Predicted probability of overweight and obesity across the percentage of days a guideline was met Note: Individual circles represtent each child with the linear predicted probability and 95% Confidence Interval

**Table 3.** Prevalence of meeting movement guidelines based on “percent days” criteria (DAYS-24hr).

| **Percent Days** | **Physical Activity**<br>Percent (%) | **Sleep**<br>Percent (%) | **Screentime**<br>Percent (%) | **All Three**<br>Percent (%) |
| --- | --- | --- | --- | --- |
| 10% | 91.4 | 99.4 | 87.0 | 63.5 |
| 20% | 83.4 | 94.9 | 78.2 | 42.9 |
| 30% | 74.9 | 88.4 | 71.0 | 29.1 |
| 40% | 66.3 | 77.9 | 62.0 | 20.2 |
| 50% | 60.6 | 66.6 | 52.9 | 13.8 |
| 60% | 49.6 | 53.6 | 41.5 | 6.9 |
| 70% | 40.9 | 40.8 | 33.1 | 3.5 |
| 80% | 32.6 | 30.6 | 26.4 | 1.7 |
| 90% | 20.7 | 15.9 | 15.3 | 0.4 |
| 100% | 8.8 | 5.7 | 6.8 | 0.2 |

### Data Handling Strategy #3 – Random Sampling of Four Days (RAND-24hr)

The percentages of participants meeting each 24-hour movement guideline for each iteration of random sampling of four days (three weekdays, one weekend day) are presented in Table 4. When averaging percentages across all 10 iterations, 56.3% of participants met physical activity guidelines, 55.5% met sleep guidelines, 37.7% met screentime guidelines, and 16.0% met all three guidelines. Over 10 iterations of randomization (Table 5), 25.2% of participants never met MVPA guidelines, 14.8% never met sleep guidelines, 42.3% never met screentime guidelines, and 63.6% never met all three guidelines. Conversely, regardless of the randomly sampled days, 34.7% always met MVPA guidelines, 23.5% always met sleep guidelines, 20.3% always met screentime guidelines, and 3.5% always met all three guidelines. The remaining participants met guidelines on one or more iterations of randomization. Specifically, 40.1% of participants met MVPA guidelines on one or more iterations, 61.7% met sleep guidelines on one or more iterations, 37.4% met screentime guidelines on one or more iterations and 32.9% met all three guidelines on one or more iterations. Figure 6 visually summarizes these results. The odds of overweight and obesity for participants meeting guidelines across all 10 random iterations is also displayed in Figure 4 (yellow circles), alongside the odds of overweight and obesity from models using the average of total observed days and compared to other previously published estimates (green circles). No two random iterations produced identical odds and variability across estimates was observed for each movement behavior guideline.

**Figure 6.**
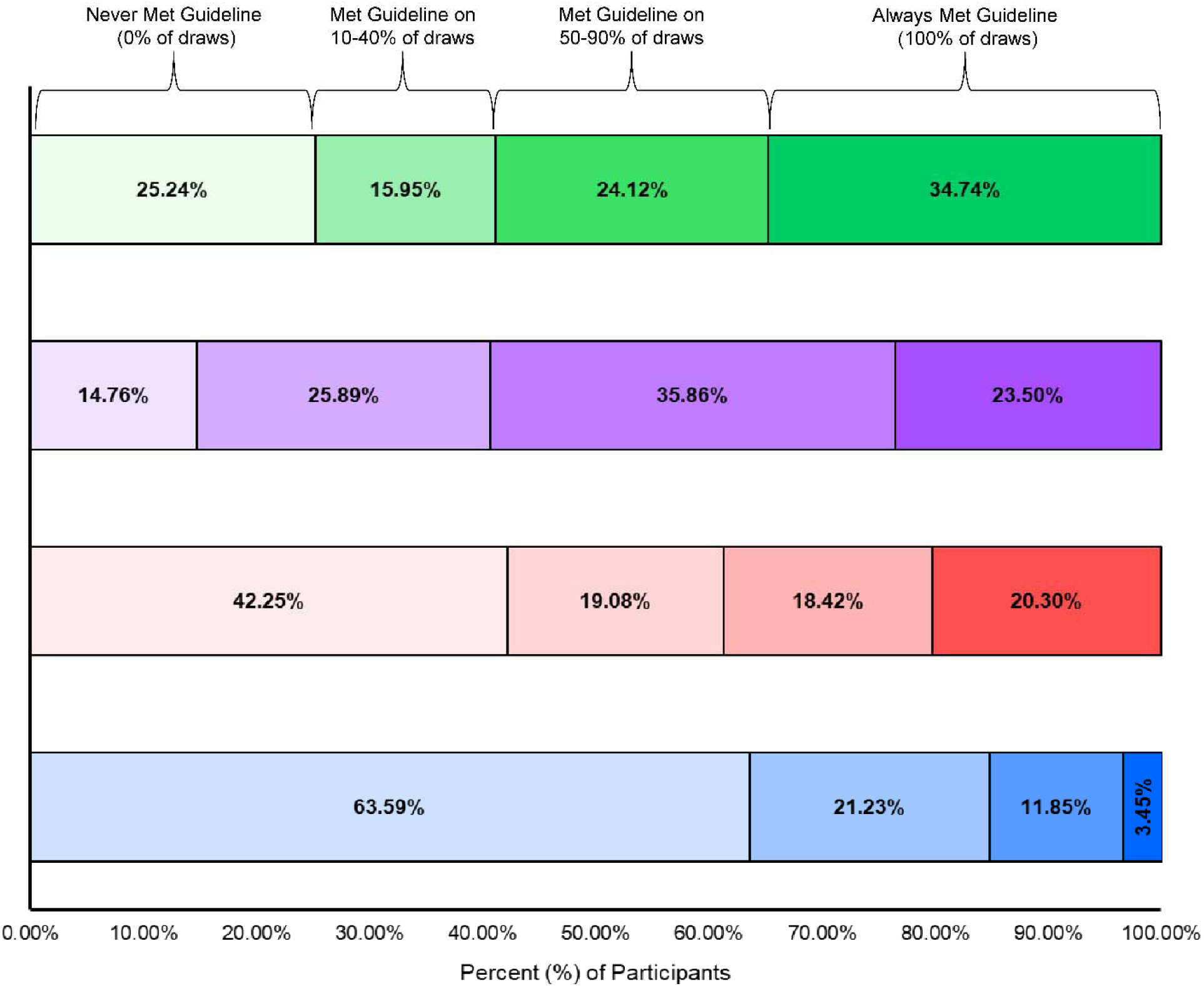
Number of times participants met guidelines after ten rounds of randomization for four days of data

**Table 4.** Classification of meeting movement guidelines based on averages from ten rounds of randomization with four days of data (AVG-24hr).

| **Randomization Round** | **Physical Activity**<br>Percent (%) | **Sleep**<br>Percent (%) | **Screentime**<br>Percent (%) | **All Three**<br>Percent (%) |
| --- | --- | --- | --- | --- |
| Round #1 | 57.1 | 54.0 | 36.9 | 16.2 |
| Round #2 | 54.9 | 55.3 | 37.2 | 15.8 |
| Round #3 | 56.4 | 57.5 | 36.4 | 15.8 |
| Round #4 | 56.8 | 54.9 | 37.5 | 16.3 |
| Round #5 | 56.0 | 57.5 | 38.6 | 15.9 |
| Round #6 | 57.3 | 55.2 | 39.0 | 16.2 |
| Round #7 | 57.5 | 55.6 | 37.4 | 16.8 |
| Round #8 | 55.1 | 55.4 | 37.1 | 15.1 |
| Round #9 | 55.4 | 55.3 | 38.9 | 16.0 |
| Round #10 | 55.5 | 54.2 | 37.8 | 15.3 |
| **Total Average** | 56.2 | 55.5 | 37.7 | 15.9 |

**Table 5.** The total number of times participants met guidelines after ten rounds of randomization for four days of data.

| **Times**<br>**Guidelines**<br>**Were Met** | **Physical Activity**<br>Percent (%) | **Sleep**<br>Percent (%) | **Screentime**<br>Percent (%) | **All Three**<br>Percent (%) |
| --- | --- | --- | --- | --- |
| 0 | 25.2 | 14.7 | 42.2 | 63.5 |
| 1 | 4.7 | 6.1 | 6.7 | 9.6 |
| 2 | 4.1 | 7.2 | 3.7 | 4.7 |
| 3 | 3.6 | 6.7 | 5.0 | 3.7 |
| 4 | 3.4 | 5.7 | 3.6 | 3.1 |
| 5 | 3.8 | 5.8 | 3.6 | 3.0 |
| 6 | 3.7 | 7.7 | 4.0 | 1.8 |
| 7 | 3.8 | 6.0 | 2.5 | 1.8 |
| 8 | 4.8 | 7.9 | 4.0 | 3.0 |
| 9 | 7.9 | 8.3 | 4.0 | 2.0 |
| 10 | 34.7 | 23.5 | 20.3 | 3.4 |

## DISCUSSION

Considerable day-to-day variability in movement behaviors was present in this cohort and applying different strategies for handling multi-day movement data (i.e., averaging across days versus classifying individual days, as meeting or not meeting guidelines) produced differing estimates of the proportion of children achieving the 24hrG and subsequent associations with overweight and obesity. Moreover, a clear dose-response relationship was observed between the number of days a child met a guideline and the probability of overweight/obesity. These findings indicate the more days a child meets the guidelines the greater the association with reduced odds of unhealthy weight. These findings have important implications for policymaking where, depending on the distillation of the 24hr data, vastly differing estimates of the prevalence of meeting the 24hrG and subsequent interpretations alongside the health outcomes of overweight and obesity may be observed. This is highlighted by the data presented where almost all children met a guideline on at least one or more days, although on average they did not meet a guideline, while conversely, the majority of children that on average met a guideline had one or more days where they did not. Thus, just because a child does not meet a 24hrG does not indicate they never meet the guideline for a given day. Based on these data, the prevalence of children who meet a 24hrG on one or more days is likely higher than what an average, which are the statistics typically reported in research and national reports, suggests.

The 24hrG are written such that each guideline should be met daily. Children should have “an accumulation of at least 60 minutes per day of moderate to vigorous physical activity”, “9 to 11 hours of sleep per night”, and “no more than 2 hours per day of screentime” on a given day.^1^ Because of this language, one could expect that a child should accumulate sufficient activity, sleep, and have reduced screentime on 100% of days. Using this interpretation of the guidelines with our current sample of children indicates only 8.8% met physical activity guidelines, 5.7% met sleep guidelines, 6.8% met screentime guidelines, and 0.2% met all three. In addition, there was a clear dose response of the percentage of days participants met guidelines and the odds of overweight and obesity, such that the odds of overweight and obesity decreased as the percentage of days needing to be classified as meeting the guideline increased. Researchers should take note of this when determining the number of days they require participants to meet the guidelines (average across days, every single day) and clearly report and justify why a particular method was chosen, as strategies can produce different results and subsequent interpretations.

A closer examination of the day-to-day estimates in Figure 1 reveals all children meet one or more guidelines on at least one day. This is further emphasized in Figures 1 and 2 where children classified, based on their average values as not meeting the guidelines, met the guidelines on some of the days. Conversely, those children classified as meeting the guidelines, based on the average values, had many days where they did not meet the guidelines. Thus, not considering day-to-day variability in movement behaviors may underrepresent the proportion of days children meet 24hrG even if their average values do not eclipse the guideline thresholds.

There is no discernible pattern common across participants in participants’ day-to-day variability of movement behaviors. On any given day, participants may meet some 24hrG while other participants fail to meet that same 24hrG, but based on an average across days, they may both meet 24hrG. Using the average of movement behaviors across sampling periods does not capture day-to-day variability of behaviors which may be important to understand. Using Figure 2 as an example, Participant #2 met MVPA guidelines on average, but did not meet the guidelines on five out of 10 days. Classification as not meeting guidelines using the average across a sampling period does not imply never meeting. Identifying day-to-day variability may be most important for those who, on average, are not meeting the guidelines, but may meet them on certain days. Using Figure 2 again for illustrative purposes, Participant #1 did not meet MVPA guidelines on average but met the guidelines on four out of 10 days. These types of issues can be observed for sleep and screentime as well. Understanding what helps a child meet a guideline on some days versus others can provide useful in designing maximally effective interventions to promote optimal movement profiles.

Variability in meeting guidelines and the odds of overweight and obesity is also highlighted for the total sample when considering the RAND-24hr data handling strategy. When tracking the total number of times participants met 24hrG across all 10 iterations of the RAND-24hr strategy, a large percentage of participants were classified as meeting guidelines during some iterations and classified as not meeting guidelines during other iterations. Specifically, 40.1% of participants were differentially classified as meeting or not meeting physical activity guidelines, 61.7% for sleep guidelines, 37.4% for screentime guidelines, and 32.9% for all three guidelines depending on which sample of four days were randomly selected. Children who comprised the sample of meeting the guidelines in each of the RAND-24hr iterations differed as well. For example, Random Iteration #1 might classify Child A as meeting all three 24hrG based on averaging the random sample of their four days of data. In Random Iteration #2, Child B might replace Child A, keeping the proportion meeting the guideline the same, but that proportion is comprised of different children from the sample. There was also variability in the odds of overweight and obesity for participants classified as meeting guidelines across all 10 iterations. Some of the random iterations demonstrated significant associations with overweight/obesity while others demonstrated non-significant findings. Thus, depending on the days sampled associations with meeting a guideline and overweight/obesity could be stronger or weaker. These nuances are important to consider when designing behavior change interventions that work on an individual-level and/or those that target behaviors and contexts on specific days. When day-specific movement behavior data are collected, we encourage researchers to report the total number of days participants met each 24hrG alongside the proportion meeting the guidelines based on the average across the days measured as well as reporting the number of days participants met all three components of the guidelines.

Having highlighted some issues that might arise when averaging movement behaviors prior to classification of meeting/not meeting the 24hrG, we acknowledge that researchers may average movement behaviors for practical reasons. For example, if movement behaviors are not measured each day and no day-specific contextual information is collected on each participant, investigating day-level associations with movement behaviors would not be feasible. An example of this is the use of a 7-day recall to measure screentime, which is common in the field.^17,28^ The 7-day recalls collect information about typical amounts of screentime each day in the past 7 days, not for each day individually. Using such a measure, it would be necessary to average physical activity and sleep data as well to ensure similar handling of the movement behaviors. Averaging movement behaviors when handling multi-day data, such as multi-day accelerometer data, is also a common convention in the field and there is a larger precedent for this type of data handling strategy.^17^ Still, averaging data allows for participants to potentially “have a day off” from meeting guidelines and making other days when they do meet the guidelines carry more weight in the average. We also realize, however, that participants may have an “off day” due to unforeseen circumstances, including illness, inclement weather, or other scenarios in which a normal day’s activities are altered. Using total averages can indeed aid in removing the bias or variability in the data brought on by these types of “off days”. Conversely, examining individual days allows for questions to be answered about day-specific predictors. Using the total number of days participants meet 24hrG could improve our understanding of the day-to-day differences of contextual influences on meeting/not meeting the guidelines. While we understand the challenges associated with collecting day-level information, these details are important to capture if we want a more granular understanding of 24hr movement behaviors among children. To address these day-specific issues, researchers are encouraged to, when possible, capture contextual information about participants’ days, which can then be used as a lens through which to interpret accelerometry-derived movement behaviors.

Previous studies examining differences in health outcomes between youth who meet guidelines on average and those who meet guidelines daily are limited. White et al.^29^ found no clinically significant differences between the cardiovascular health of participants who met PA guidelines daily and those who achieved the same weekly activity condensed into a few days. However, this study only included participants who were already meeting the PA guideline, thus limiting the sample to those who were highly active in the first place. We found a clear downward trend in the predicted probability of overweight and obesity as the percentage of days needed to meet each guideline increased. In addition, the predicted probability of overweight and obesity for participants when meeting guidelines on 100% of measured days was consistently lower when compared to predicted probabilities based on the average across all measured days for all movement behaviors. Still, future studies are needed on a wide range of health outcomes to understand how meeting 24hrG daily might differentially impact other health outcomes compared to meeting 24hrG on average.

### Strengths and Limitations

This methodological exploration of data handling strategies had several strengths. These include 1) a large sample size, 2) a relatively lengthy accelerometer data collection period, 3) a large dataset of complete activity measurements on physical activity, sleep, and screentime, and 4) being one of the first studies to explore the important methodological issue of data handling strategies and how they may differentially impact subsequent movement guideline and health outcome interpretations. Previous studies have explored this issue but have only done so with participants who have a limited number of measured days and are already meeting the movement guidelines.^29^

There are several limitations to this study as well. While the sample used to illustrate data handling strategies was representative of children in the southeastern region of the United States, results may not apply to researchers working with older populations. We believe reporting the total number of days participants met guidelines alongside averages is still good practice, regardless of the participants’ age. The specific data handling strategies that were chosen for this study may not encompass all data handling strategies that are currently being employed in the field. The choice to use a sample of four days (including three weekdays and one weekend day) was based off what has been commonly done in the past with studies that measure children’s physical activity and sleep via accelerometry and screentime with questionnaires.^17^ In terms of the sleep guidelines, we did not explore bed/wake time consistency as this is not a commonly investigated outcome in the 24hrG literature, but is part of the guidelines. Most studies utilizing the 24hrG as an outcome of interest focus on total sleep time estimates, but future studies should consider investigating bed/wake time consistency and how data handling strategies may differentially influence interpretations of this data. For screentime, we utilized a parent-reported measure, which was reported in increments of 30 minutes. Because of the 30-minute increment, parents may have been forced to under or over report screentime for their child on a given day. While self or parent-reported screentime is commonly used at various levels of granularity (10, 15, 20, 30-minute increments), this is a methodological weakness of our data and may influence the overall screentime results and the variability of the screentime data as well.

## CONCLUSION

Different data handling strategies produce varying estimates of children meeting the 24-hour movement guidelines and associations with overweight and obesity. Not accounting for day-to-day variability in children’s movement behaviors and using a limited sample of measured days may produce lower or higher estimates of children meeting physical activity, sleep, and screentime guidelines and may influence health outcome results that are statistically linked to meeting or not meeting the guidelines. Furthermore, aggregating data across days limits the understanding of factors associated with achieving one or more 24hrG for a given day. Such data could provide insights into factors influencing meeting the 24hrGs which could be targeted for public health interventions.

We recommend researchers who use 24hrG, whether as predictors or outcomes, should utilize the distillation procedures used in this study and report the total number of days participants meet 24hrG and use them as part of analyses to understand day-to-day variability in movement behaviors as well. Utilizing daily estimates of meeting 24hrG may also help identify important day-level contextual factors that could be harnessed within an intervention to promote adherence to 24hrG and help researchers further refine our understanding of the dose-response relationship of meeting 24hrG and health outcomes. Finally, researchers should be aware of how data handling strategies prior to classification might impact estimates regarding 24hrG and health outcomes and should clearly report how movement data was handled prior to classification, regardless of the strategies used.

## LIST OF ABBREVIATIONS

24hrG: 24-hour movement behavior guidelines
AVG-24hr: Data handling strategy in which estimates were averaged across the entire measurement period
DAYS-24hr: Data handling strategy in which each individual day was classified as meeting/not meeting guidelines
MVPA: Moderate to Vigorous Physical Activity
PA: Physical Activity
RAND-24hr: Data handling strategy in which a random sample of four days was used to classify guideline adherence across 10 iterations.

## DECLARATIONS

### Ethics approval and consent to participate

All procedures were approved by the University of South Carolina’s Institutional Review Board prior to the start of the study (IRB#Pro00080382) and participant consent was obtained prior to being enrolled in the study.

### Consent for publication

Not applicable.

### Availability of data and material

The datasets used for the current study are available from the corresponding author on reasonable request.

### Authors’ contributions

Conceptualization: CDP, SB, and MWB

Methodology: CDP, SB, and MWB

Formal analysis: CDP and MWB

Data curation: CDP, SB, MWB, RD, HP, and LV

Writing – Original Draft: CDP, SB, MWB, RD, HP, LV, ADO, and RGW

Writing – Review and Editing: CDP, SB, MWB, RD, HP, LV, ADO, and RGW

Visualization: CDP, SB, RGW, and MWB

Funding acquisition: MWB, SB, RD, and LV

All authors read and approved the final manuscript.

## Acknowledgements

Not applicable.

## REFERENCES

1. Tremblay MS, Carson V, Chaput JP, et al. Canadian 24-Hour Movement Guidelines for Children and Youth: An Integration of Physical Activity, Sedentary Behaviour, and Sleep. Appl Physiol Nutr Me. Jun 2016;41(6):S311–S327. doi:10.1139/apnm-2016-0151

2. Tremblay MS, Ross R. How should we move for health? The case for the 24-hour movement paradigm. Can Med Assoc J. Dec 7 2020;192(49):E1728–E1729. doi:10.1503/cmaj.202345

3. Okely AD, Ghersi D, Loughran SP, et al. A collaborative approach to adopting/adapting guidelines. The Australian 24-hour movement guidelines for children (5-12 years) and young people (13-17 years): An integration of physical activity, sedentary behaviour, and sleep. Int J Behav Nutr Phys Act. Jan 6 2022;19(1):2. doi:10.1186/s12966-021-01236-2

4. Willumsen J, Bull F. Development of WHO Guidelines on Physical Activity, Sedentary Behavior, and Sleep for Children Less Than 5 Years of Age. J Phys Act Health. Jan 2020;17(1):96–100. doi:10.1123/jpah.2019-0457

5. Draper CE, Tomaz SA, Biersteker L, et al. The South African 24-Hour Movement Guidelines for birth to 5 years: An integration of physical activity, sitting behavior, screen time, and sleep (vol 17, pg 109, 2020). J Phys Act Health. Apr 2020;17(4):491–491. doi:10.1123/jpah.2020-0067

6. Feng J, Zheng C, Sit CHP, Reilly JJ, Huang WY. Associations between meeting 24-hour movement guidelines and health in the early years: A systematic review and meta-analysis. J Sport Sci. Nov 17 2021;39(22):2545–2557. doi:10.1080/02640414.2021.1945183

7. Roman-Vinas B, Chaput JP, Katzmarzyk PT, et al. Proportion of children meeting recommendations for 24-hour movement guidelines and associations with adiposity in a 12-country study. Int J Behav Nutr Phys Act. Nov 25 2016;13(1):123. doi:10.1186/s12966-016-0449-8

8. Lemos L, Clark C, Brand C, et al. 24-hour movement behaviors and fitness in preschoolers: A compositional and isotemporal reallocation analysis. Scand J Med Sci Sports. Jun 2021;31(6):1371–1379. doi:10.1111/sms.13938

9. Khan A, Lee EY, Tremblay MS. Meeting 24-h movement guidelines and associations with health related quality of life of Australian adolescents. J Sci Med Sport. May 2021;24(5):468–473. doi:10.1016/j.jsams.2020.10.017

10. Xiong XQ, Dalziel K, Carvalho N, Xu RB, Huang L. Association between 24-hour movement behaviors and health-related quality of life in children. Qual Life Res. Jan 2022;31(1):231–240. doi:10.1007/s11136-021-02901-6

11. Sampasa-Kanyinga H, Chaput JP, Goldfield GS, et al. 24-hour movement guidelines and suicidality among adolescents. J Affect Disorders. Sep 1 2020;274:372–380. doi:10.1016/j.jad.2020.05.096

12. Sampasa-Kanyinga H, Lien A, Hamilton HA, Chaput JP. The Canadian 24-hour movement guidelines and self-rated physical and mental health among adolescents. Can J Public Health. Apr 2022;113(2):312–321. doi:10.17269/s41997-021-00568-7

13. Thivel D, Tremblay MS, Katzmarzyk PT, et al. Associations between meeting combinations of 24-hour movement recommendations and dietary patterns of children: A 12-country study. Prev Med. Jan 2019;118:159–165. doi:10.1016/j.ypmed.2018.10.025

14. Cliff DP, McNeill J, Vella SA, et al. Adherence to 24-Hour Movement Guidelines for the Early Years and associations with social-cognitive development among Australian preschool children. Bmc Public Health. Nov 20 2017;17doi:ARTN 85710.1186/s12889-017-4858-7

15. Walsh JJ, Barnes JD, Cameron JD, et al. Associations between 24 hour movement behaviours and global cognition in US children: a cross-sectional observational study. Lancet Child Adolesc. Nov 2018;2(11):783–791. doi:10.1016/S2352-4642(18)30278-5

16. Taylor RW, Haszard JJ, Meredith-Jones KA, et al. 24-h movement behaviors from infancy to preschool: cross-sectional and longitudinal relationships with body composition and bone health. Int J Behav Nutr Phy. Nov 26 2018;15doi:ARTN 11810.1186/s12966-018-0753-6

17. Tapia-Serrano M, Sevil-Serrano, JS, Sanchez-Miguel, PA, Lopez-Gil, JF, Tremblay, MS, Garcia-Hermoso, A. Prevalence of meeting 24-Hour Movement Guidelines from pre-school to adolescence: A systematic review and meta-analysis including 387,437 participants and 23 countries. Journal of Sport and Health Science. 2022;10.1016/j.jshs.2022.01.005

18. Vale S, Silva P, Santos R, Soares-Miranda L, Mota J. Compliance with physical activity guidelines in preschool children. J Sports Sci. Apr 2010;28(6):603–8. doi:10.1080/02640411003702694

19. Beets MW, Bornstein D, Dowda M, Pate RR. Compliance with national guidelines for physical activity in U.S. preschoolers: measurement and interpretation. Pediatrics. Apr 2011;127(4):658–64. doi:10.1542/peds.2010-2021

20. Kharlova I, Fredriksen MV, Mamen A, Fredriksen PM. Daily and Weekly Variation in Children’s Physical Activity in Norway: A Cross-Sectional Study of The Health Oriented Pedagogical Project (HOPP). Sports (Basel). Nov 20 2020;8(11)doi:10.3390/sports8110150

21. Ridgers ND, Barnett LM, Lubans DR, Timperio A, Cerin E, Salmon J. Potential moderators of day-to-day variability in children’s physical activity patterns. J Sports Sci. Mar 2018;36(6):637–644. doi:10.1080/02640414.2017.1328126

22. Becker SP, Sidol CA, Van Dyk TR, Epstein JN, Beebe DW. Intraindividual variability of sleep/wake patterns in relation to child and adolescent functioning: A systematic review. Sleep Med Rev. Aug 2017;34:94–121. doi:10.1016/j.smrv.2016.07.004

23. Bei B, Wiley JF, Trinder J, Manber R. Beyond the mean: A systematic review on the correlates of daily intraindividual variability of sleep/wake patterns. Sleep Med Rev. Aug 2016;28:108–24. doi:10.1016/j.smrv.2015.06.003

24. Nicholson L, Bohnert AM, Crowley SJ. A developmental perspective on sleep consistency: Preschool age through emerging adulthood. Behav Sleep Med. Jan 11 2022:1–20. doi:10.1080/15402002.2021.2024192

25. Migueles J, Rowlands, AV, Huber, F, Sabia, Severine, van Hees, VT. GGIR: A Research Community–Driven Open Source R Package for Generating Physical Activity and Sleep Outcomes From Multi-Day Raw Accelerometer Data. Journal for the Measurement of Physical Behaviour. 2019;2(3):188–196. 10.1123/jmpb.2018-0063

26. Hildebrand M, Hansen BH, van Hees VT, Ekelund U. Evaluation of raw acceleration sedentary thresholds in children and adults. Scand J Med Sci Spor. Dec 2017;27(12):1814–1823. doi:10.1111/sms.12795

27. van Hees VT, Sabia S, Anderson KN, et al. A Novel, Open Access Method to Assess Sleep Duration Using a Wrist-Worn Accelerometer. Plos One. Nov 16 2015;10(11)doi:ARTN e014253310.1371/journal.pone.0142533

28. Fang K, Mu M, Liu K, He Y. Screen time and childhood overweight/obesity: A systematic review and meta-analysis. Child Care Health Dev. Sep 2019;45(5):744–753. doi:10.1111/cch.12701

29. White DA, Willis EA, Ptomey LT, Gorczyca AM, Donnelly JE. Weekly Frequency of Meeting the Physical Activity Guidelines and Cardiometabolic Health in Children and Adolescents. Med Sci Sports Exerc. Jan 1 2022;54(1):106–112. doi:10.1249/MSS.0000000000002767

